# Protection and waning of natural and hybrid COVID-19 immunity

**DOI:** 10.1101/2021.12.04.21267114

**Authors:** Yair Goldberg, Micha Mandel, Yinon M. Bar-On, Omri Bodenheimer, Laurence Freedman, Nachman Ash, Sharon Alroy-Preis, Amit Huppert, Ron Milo

**Affiliations:** Technion - Israel Institute of Technology, Israel; The Hebrew University of Jerusalem, Israel; Department of Plant and Environmental Sciences, Weizmann Institute of Science, Israel; Israel Ministry of Health, Israel; The Bio-statistical and Bio-mathematical Unit, The Gertner Institute for Epidemiology & Health Policy Research, Sheba Medical Center, Israel; The Sackler Faculty of Medicine, Tel Aviv University, Israel

## Abstract

**BACKGROUND:** Infection with SARS-CoV-2 provides substantial natural immunity against reinfection. Recent studies have shown strong waning of the immunity provided by the BNT162b2 vaccine. The time course of natural and hybrid immunity is unknown.

**METHODS:** Data on confirmed SARS-CoV-2 infections were extracted from the Israeli Ministry of Health database for the period August to September 2021 regarding all persons previously infected or vaccinated. We compared infection rates as a function of time since the last immunity-conferring event using Poisson regression, adjusting for possible confounding factors.

**RESULTS:** Confirmed infection rates increased according to time elapsed since the last immunity-conferring event in all cohorts. For unvaccinated previously infected individuals they increased from 10.5 per 100,000 risk-days for those previously infected 4-6 months ago to 30.2 for those previously infected over a year ago. For individuals receiving a single dose following prior infection they increased from 3.7 per 100,000 person days among those vaccinated in the past two months to 11.6 for those vaccinated over 6 months ago. For vaccinated previously uninfected individuals the rate per 100,000 person days increased from 21.1 for persons vaccinated within the first two months to 88.9 for those vaccinated more than 6 months ago.

**CONCLUSIONS:** Protection from reinfection decreases with time since previous infection, but is, nevertheless, higher than that conferred by vaccination with two doses at a similar time since the last immunity-conferring event. A single vaccine dose after infection helps to restore protection.

## Introduction

While a rapid decline in protection against SARS-CoV-2 after two doses of BNT162b2 vaccination has been observed in several studies,^1–3^ the levels of protection and the presence or extent of waning of natural immunity are still unclear. Several studies have reported a substantial natural immunity six and more months following infection,^4–8^ although one recent study^9^ reported mRNA based vaccines to have a 5-fold higher protection against hospitalization compared to protection provided by prior infection. Waning in both the humoral and cellular responses of the immune system is well documented in vaccinated and in previously infected persons.^10,11^ In addition, studies of seasonal corona viruses have demonstrated waning of natural immunity.^12^ It is also unclear how natural immunity interacts with immunity conferred by vaccination. Some laboratory studies have suggested that “hybrid immunity” (immunity conferred by previous infection combined with vaccination) elicits neutralizing antibodies at higher levels,^13^ that it is more broad-spectrum^14^, and that it provides more protection against infection^15^ than immunity conferred by vaccination or infection alone. The durability of immunity resulting from infection with SARS-CoV-2, and how it compares to that of vaccination is an essential question at both the individual and national levels.

We study the time course of natural immunity resulting from infection by estimating the rates of confirmed SARS-CoV-2 infection among previously infected, unvaccinated individuals, previously infected individuals who have also received the BNT162b2 vaccine, and vaccinated individuals without previous infection, according to the time elapsed since infection or vaccination. We quantify, within these groups, the association of time since infection or vaccination with the rate of confirmed infection. We also compare rates of infection between these groups, allowing assessment of the level of protection afforded by “hybrid immunity” relative to natural immunity or to immunity conferred by vaccination.

## Methods

The analysis is based on the Israel Ministry of Health’s database. Israel has experienced four pandemic waves, with very rapid vaccination campaigns offering two doses of the BNT162b2 vaccine and a third booster dose (see Supplementary Appendix). In March 2021, previously infected individuals were eligible to receive a single BNT162b2 dose at least three months after recovery from Covid-19. In this study, re-infection is defined as a positive PCR test in an individual who had a previous positive result on a sample taken at least 90 days earlier.^16^ Severe disease is defined following the US NIH definition: resting respiratory rate of more than 30 breaths per minute, an oxygen saturation of less than 94% while breathing ambient air, or a ratio of partial pressure of arterial oxygen to fraction of inspired oxygen of less than 300. The MoH database also includes basic demographic information, such as sex, age, place of residency, and population sector.

We study rates of confirmed infections over the study period August 1 to September 30, 2021, during which the Delta variant was dominant in Israel.^17^ We extracted personal data from the MoH database on all individuals aged 16 or above who had tested positive before July 1, 2021, or who had received at least two BNT162b2 vaccination doses at least 7 days before the end of the study period. We excluded from the analysis individuals who had missing data on age group or gender, those who tested positive between July 1 and July 31, 2021 (the day before the start of the study period), those who had recovered from a PCR-confirmed Covid-19 infection and then received more than one BNT162b2 dose (a small group with limited follow-up data), those who had stayed abroad during the whole study period, and those who vaccinated with a vaccine different than BNT162b2 before August 1; see Figure 1 for details. We compared incidence rates over the study period among individuals with different histories of immunity-conferring events:

**Figure 1.**
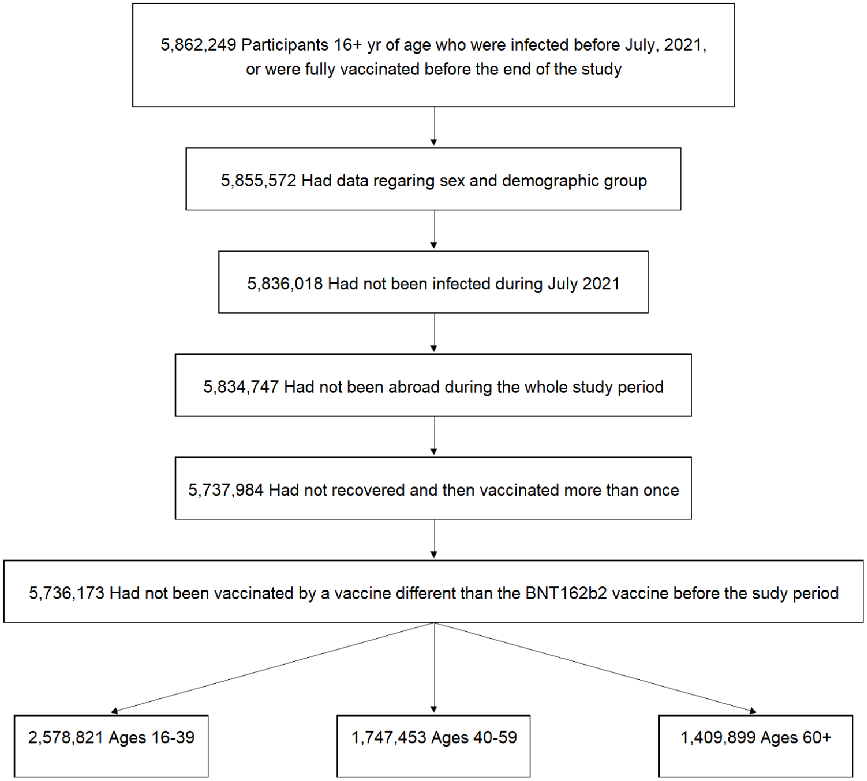
Study population. The participants in the study included persons who were 16 years of age or older, who were infected by SARS-CoV-2 before July 1, 2021 or were doubly-vaccinated, had available data regarding sex and demographic sector, had no documented positive PCR result between July 1, 2021 and July 30, 2021, had not stayed abroad during the whole study period, had received at most one vaccine dose after recovery, and had not been vaccinated with a vaccine different fromBNT162b2 before August 1. Age groups as of January 1, 2021.

- **Recovered:** Previously infected individuals 90 or more days after confirmed infection who had never been vaccinated.
- **Recovered then Vaccinated:** Previously infected individuals who later were 7 or more days after receiving a single vaccine dose.
- **Vaccinated then Recovered:** Individuals who had been vaccinated with one or two doses and were later infected.
- **Vaccinated:** Individuals seven days or more after receiving the second dose, and who had not been infected before the start of the study period.
- **Booster:** Individuals who received a third (booster) dose 12 or more days previously and had not been infected before the start of the study period.

These cohorts were divided into sub-cohorts according to the time elapsed from the last immunity-conferring event. We used two-month periods as our basic time interval for defining the sub-cohorts, but combined months 12 to18 for the Recovered cohort and omitted months 8 to 10 for the Vaccinated and the hybrid cohorts due to the small number of individuals. An individual could contribute follow-up days to different sub-cohorts, and could also move between cohorts according to the following rules. A recovered individual who, during the study period, received a first BNT162b2 dose exited the Recovered cohort on the day of vaccination and entered the Recovered then Vaccinated cohort seven days later. A recovered individual who had received a first vaccine dose but then received a second dose during the study period exited the Recovered then Vaccinated cohort at the time of the second vaccination. An individual in the Vaccinated cohort who received the booster dose during the study period, exited that cohort on the day of the booster dose and entered the Booster cohort 12 days later.^18^ A person who was infected between May 1 and June 30, 2021 entered the Recovered or Recovered then Vaccinated cohort (according to previous vaccination status) 90 days after the positive test. A person who received a vaccine different from BNT162b2 exited his cohort on the day of vaccination.

Typically, infection rates among recovered or vaccinated individuals are compared to the infection rate among unvaccinated-not-previously-infected persons. However, due to the high vaccination rate in Israel, the latter cohort is small and unrepresentative of the overall population; furthermore, the MoH database does not include complete information on such individuals. Therefore, we did not include unvaccinated-not-previously-infected individuals in the analysis.

We analyzed the data using a methodology similar to that used in our previous studies.^8,18,19^ The number of confirmed infections and the number of days at risk during the study period were counted for each sub-cohort. A Poisson regression model was fitted adjusting for age group (16-39, 40-59, and 60+ years; age as of January 1, 2021), sex, population group (General Jewish, Arab, ultra-Orthodox Jewish), calendar week, and an exposure risk measure. The latter was calculated for each person on each follow-up day according to the proportion of new confirmed infections during the past seven days in their area of residence; this continuous measure was then divided into ten categories according to deciles (see Bar-On et al.^18^ for details). An average risk was imputed to individuals with missing data on residency. A model with an interaction between age group and sub-cohort was also fitted in order to estimate age-specific incidence rates for each sub-cohort. In case of infection, an individual contributed an event to their current sub-cohort. Based on the estimated parameters of the fitted regression model, the incidence rate of each sub-cohort, adjusted for the confounders, was calculated as the expected number of events if all days-at-risk were in that sub-cohort, and is presented per 100,000 days (see Supplementary Appendix). Confidence intervals (95% level) were calculated using a bootstrap-like simulation approach^20^ without an adjustment for multiplicity. There were too few cases for an in-depth comparison of severe disease, so only a descriptive analysis was performed.

## Results

### Study Population and Descriptive Statistics

The analysis is based on more than 5.7 million individuals who contributed days to the five main cohorts (Figure 1). Figure 2 shows the dynamics of the cohorts over time, with the area under the lines over the study period representing the number of person-days at risk for each cohort. Table 1 presents the number of events (confirmed infections and severe disease) by cohorts and demographics, and shows the distribution of person-days at risk by gender, age and population group in the different cohorts. The gender distribution was similar among the five cohorts, with only slightly more person-days for women than for men. There was a clear difference between the cohorts in the distribution of the other covariates; while 53.4% of person-days in the Booster cohort were of persons aged 60 or older, only 8.3% of the Recovered cohort, 13.8% of the Recovered then Vaccinated, 22.1% of the Vaccinated then Recovered, and 12.6% of the Vaccinated cohort person-days were from this age group. The distributions of person-days by population group also differed between the cohorts, reflecting the fact that the Arab and ultra-Orthodox Jewish groups experienced higher incidence rates of infections during the pandemic, resulting in higher proportions of these groups in the cohorts of recovered persons than in the cohorts of persons not previously infected.

**Table 1:** Demographic and clinical characteristics of the different cohorts.The table presents the proportion of person-days at risk instead of individuals as people can move between cohorts. Only person-days and events that were used in the main analysis are presented. Data are for the study period August 1, 2021, to September 30, 2021.

| Group | Recovered<br>Person-days at risk =<br>17,880,995 |  |  | Booster<br>Person-days at risk =<br>80,428,946 |  |  | Vaccinated<br>Person-days at risk =<br>184,214,308 |  |  | Recovered then Vaccinated<br>Person-days at risk =<br>9,670,155 |  |  | Vaccinated then Recovered<br>Person-days at risk =<br>1,968,832 |  |  |
| --- | --- | --- | --- | --- | --- | --- | --- | --- | --- | --- | --- | --- | --- | --- | --- |
|  | %<br>person<br>days at<br>risk | #<br>infections | #<br>severe<br>Covid-19 | %<br>person<br>days at<br>risk | #<br>infections | #<br>severe<br>Covid-19 | %<br>person<br>days at<br>risk | #<br>infections | #<br>severe<br>Covid-19 | %<br>person<br>days at<br>risk | #<br>infections | #<br>severe<br>Covid-19 | %<br>person<br>days at<br>risk | #<br>infections | #<br>severe<br>Covid-19 |
| Female | 53.1% | 2,370 | 9 | 51.0% | 2,835 | 70 | 51.2% | 77,475 | 579 | 51.7% | 441 | 8 | 51.8% | 177 | 2 |
| Male | 46.9% | 2,021 | 16 | 49.0% | 3,010 | 108 | 48.8% | 63,003 | 793 | 48.3% | 381 | 5 | 48.2% | 166 | 3 |
| Age 16-39 | 66.7% | 3,361 | 6 | 16.4% | 1,156 | 1 | 56.7% | 84,471 | 44 | 57.2% | 543 | 1 | 43.8% | 197 | 0 |
| Age 40-59 | 25.0% | 915 | 10 | 30.1% | 2,042 | 13 | 30.7% | 42,825 | 264 | 29.1% | 209 | 5 | 34.1% | 106 | 0 |
| Age 60+ | 8.3% | 115 | 9 | 53.4% | 2,647 | 164 | 12.6% | 13,182 | 1,064 | 13.8% | 70 | 7 | 22.1% | 40 | 5 |
| General Jewish | 50.7% | 2,522 | 12 | 89.2% | 4,957 | 155 | 74.1% | 111,174 | 1,071 | 54.4% | 539 | 8 | 65.9% | 255 | 5 |
| Arab | 28.4% | 1,463 | 4 | 4.1% | 417 | 12 | 5.6% | 12,328 | 74 | 22.1% | 170 | 2 | 13.5% | 50 | 0 |
| Ultra-Orthodox | 20.9% | 406 | 9 | 6.7% | 471 | 11 | 20.3% | 16,976 | 227 | 23.6% | 113 | 3 | 20.6% | 38 | 0 |

**Figure 2.**
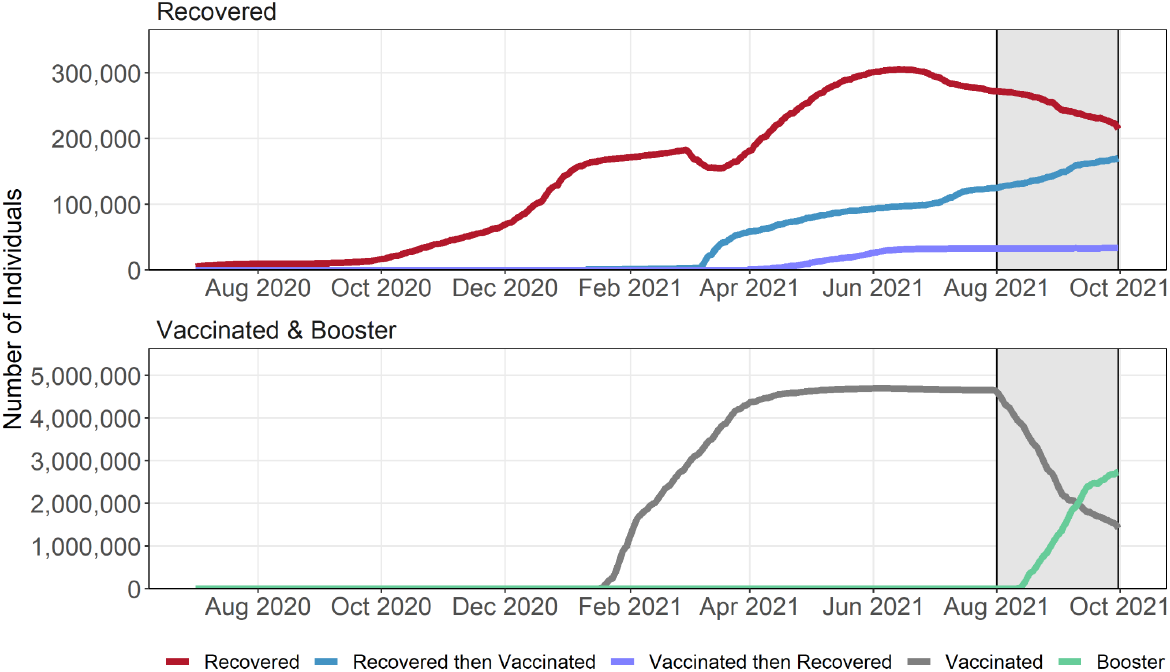
Dynamics of the cohorts. The lines represent the number of individuals belonging to each cohort over time. The numbers increase and decrease as individuals join and exit the cohorts. The shaded area indicates the study period ((August 1, 2021 to September 30, 2021).

Tables S1-S4 in the Supplementary Appendix provide a more detailed tabulation of the data, with each cohort divided into sub-cohorts according to time elapsed since infection or vaccination. As expected, the differences in the distributions of covariates between the sub-cohorts of each cohort were smaller than those between the cohorts. The most prominent differences between sub-cohorts were related to older individuals tending to receive vaccination earlier (according to the Israeli vaccination prioritization schedule). The number of person-days for sub-cohorts of recovered individuals, vaccinated or not, were much smaller than those in the Vaccinated and Booster sub-cohorts. The numbers of severe Covid-19 cases among individuals in the recovered sub-cohorts were small (less than ten), precluding reliable quantification of their levels of protection against severe disease. We therefore focused on comparing the rates of confirmed infection among the sub-cohorts.

### Waning Immunity Against Reinfection

Table 2 summarizes the results of the Poisson regression analysis, showing the incidence rate of confirmed infection per 100,000 person-days in each sub-cohort, adjusted for age, gender, population group and risk of exposure. Table 2 also provides the rate ratios for each sub-cohort, relative to the reference sub-cohort of previously uninfected individuals for whom time from vaccination was 2 months or less. The complete set of parameter estimates of the regression model is provided in Table S5 of the Supplementary Appendix. Figure 3 presents the adjusted incidence rates according to sub-cohort during the study period. The adjusted incidence rates within age groups (16-39, 40-59, and 60+) are provided in Table S6 and Figure S1 of the Supplementary Appendix.

**Table 2.** Summary of the results regarding confirmed infections of the Poisson regression analysis for all sub-cohorts. For each sub-cohort, the table shows the estimated covariate-adjusted (to the Israeli population during the study period, August 1, 2021, to September 30, 2021) confirmed infection rates per 100,000 person days, as well as the rate ratio of confirmed infections between individuals with a fresh second dose vaccination (up to two months) who were not previously infected relative to each of the other sub-cohorts. 95% confidence intervals without adjustment for multiplicity are given in square brackets. The Recovered 12+ sub-cohort includes the period 12-18 months.

| Sub-cohort | Adjusted Confirmed Infection Rate Per 100,000* | Rate ratio of reference relative to sub-cohort |
| --- | --- | --- |
| Recovered 4-6 | 10.5 [8.8, 12.4] | 2.0 [1.7, 2.4] |
| Recovered 6-8 | 14.0 [13.3, 14.8] | 1.5 [1.4, 1.6] |
| Recovered 8-10 | 20.6 [19.2, 22.1] | 1.0 [0.9, 1.1] |
| Recovered 10-12 | 28.5 [26.9, 30.2] | 0.7 [0.7, 0.8] |
| Recovered 12+ | 30.2 [28.5, 32] | 0.7 [0.6, 0.8] |
| Booster 0-2 | 8.2 [8.0, 8.5] | 2.6 [2.4, 2.7] |
| Vaccinated 0-2 | 21.1 [20, 22.4] | Reference |
| Vaccinated 2-4 | 45.0 [43.7, 46.4] | 0.5 [0.4, 0.5] |
| Vaccinated 4-6 | 69.2 [68.6, 69.8] | 0.3 [0.3, 0.3] |
| Vaccinated 6-8 | 88.9 [88.3, 89.6] | 0.2 [0.2, 0.3] |
| Recovered then Vaccinated 0-2 | 3.7 [3.1, 4.5] | 5.7 [4.6, 6.9] |
| Recovered then Vaccinated 2-4 | 4.3 [3.5, 5.2] | 5.0 [4.0, 6.1] |
| Recovered then Vaccinated 4-6 | 10.3 [9.4, 11.4] | 2.0 [1.8, 2.3] |
| Recovered then Vaccinated 6-8 | 11.6 [10.0, 13.5] | 1.8 [1.5, 2.1] |
| Vaccinated then Recovered 4-6 | 12.8 [9.9, 16.6] | 1.7 [1.3, 2.1] |
| Vaccinated then Recovered 6-8 | 17.2 [15.2, 19.2] | 1.2 [1.1, 1.4] |

\* Adjusted for age, sex, population group, calendar week, and risk of exposure

**Figure 3:**
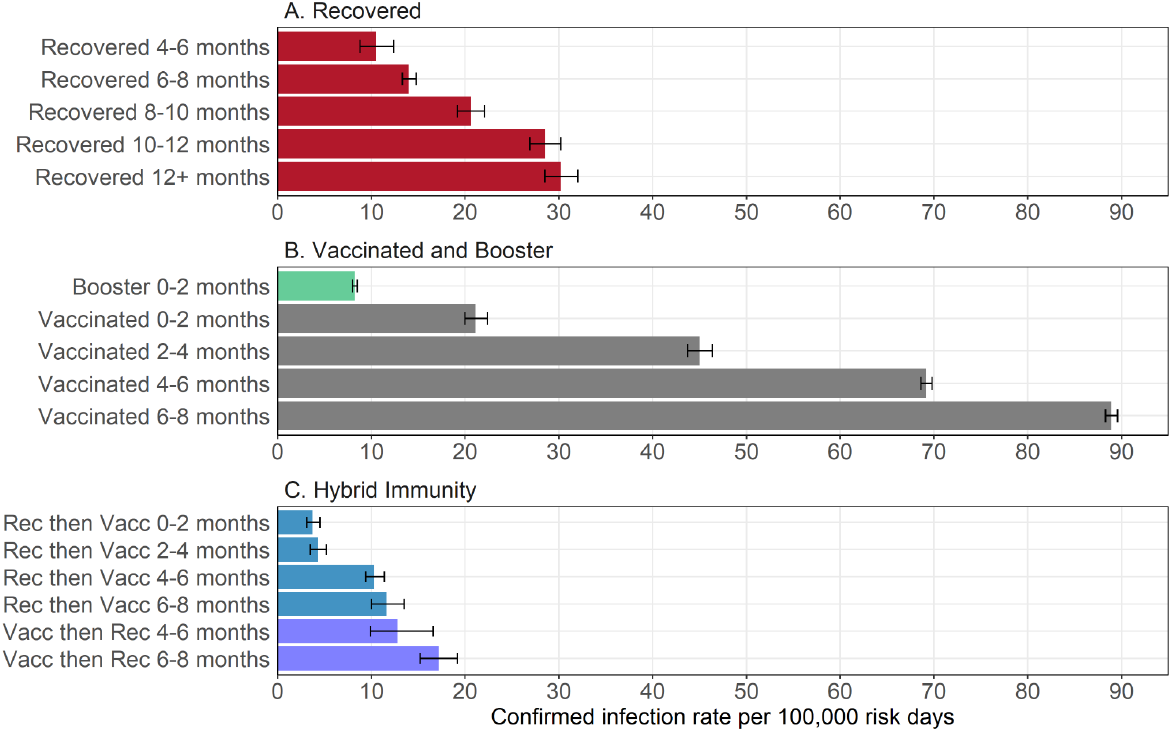
Estimated covariate-adjusted rates of confirmed infections per 100,000 at-risk days obtained from the Poisson regression analysis for the study period August 1, 2021, to September 30, 2021, stratified by sub-cohorts. Confidence intervals are not adjusted for multiplicity.

Clear evidence of waning immunity is evident for all cohorts. The rate of confirmed infections for Recovered individuals for whom the time elapsed from infection was 4 to 6 months was 10.5 per 100,000 person days (95 CI: 8.8 to 12.4) increasing with time since recovery to 30.2 (95% CI: 28.5 to 32.0) at more than 12 months. For the Vaccinated cohort, the rates were 21.1 (95% CI: 20.0 to 22.4) when the time since vaccination was less than two months increasing with time since vaccination to 88.9 (95% CI: 88.3 to 89.6) at 6 to 8 months. For the Recovered then Vaccinated cohort with the same times since vaccination, the rates were 3.7 (95% CI: 3.1 to 4.5) and 11.6 (95% CI: 10.0 to 13.5), respectively.

The adjusted rates of confirmed infection for the sub-cohorts of Recovered individuals were similar to the rates of the Recovered then Vaccinated and Vaccinated then Recovered sub-cohorts when the time elapsed since the last immunity-conferring event (either infection or vaccination) was the same (see Figure 3). For example, when the time from the last immunity-conferring event was 4 to 6 months, the rates per 100,000 days at risk were 10.5 (95% CI: 8.8 to 12.4), 10.3 (95% CI: 9.4 to 11.4), and 12.8 (95% CI: 9.9 to 16.6) for the Recovered, the Recovered then Vaccinated, and the Vaccinated then Recovered cohorts, respectively. When the time from the last immunity-conferring event was 6 to 8 months, the rates were 14.0 (95% CI: 13.3 to 14.8), 11.6 (95% CI: 10.0 to 13.5), and 17.2 (95% CI: 15.2 to 19.2) respectively. These rates are lower than the rates of doubly-vaccinated individuals, with 69.2 (95% CI: 68.8 to 69.8) 4 to 6 months after second dose and 88.9 (95% CI: 88.3 to 89.6) after 6 to 8 months, but the protection can be resorted by administration of a booster showing an adjusted rate of 8.2 (95% CI: 8.0 to 8.5) when time since the booster is less than two months (see Table 2).

### Analysis of Severe Cases

The number of severe cases among infected individuals was relatively small in most cohorts, with 25 among Recovered, 13 among Recovered then Vaccinated, and 5 among Vaccinated then Recovered, 1,372 among the Vaccinated, and 178 among the Booster individuals (Table 1). The resulting crude rates of severe disease for persons age 60 or older, ignoring the time from the last immunity-conferring event, were 0.6 per 100,000 person days for the Recovered cohort, 0.5 for the Recovered then Vaccinated cohorts, 1.1 for the Vaccinated then Recovered, 4.6 for the Vaccinated cohort, and only 0.4 for the Booster.

## Discussion

We have evaluated the level of protection against infection with SARS-Cov-2 among individuals who recovered from a previous infection and among previously uninfected individuals who received the BNT162b2 vaccine, and have studied how these levels of protection decrease over time. We compared these groups to individuals who were vaccinated and later infected with SARS-Cov-2 and to individuals who recovered from SARS-Cov-2 infection and later received a single dose of vaccine. Previous studies demonstrated the relatively higher protection of previously-infected individuals with or without an additional vaccination dose compared to previously uninfected doubly-vaccinated individuals with mRNA vaccines.^6,7^ This study is the first to comprehensively quantify the waning of natural and hybrid immunity at the national level in a real world setting.

Clear evidence of waning immunity is evident for the Recovered cohort (Figure 3A), the Vaccinated cohort (Figure 3B), and the Hybrid immunity cohorts (Figure 3C). This pattern of waning immunity was evident across all the age groups (Table S6 and Figure S1 of the Supplementary Appendix). The adjusted rates of confirmed infection for the Recovered sub-cohorts were lower than those of Vaccinated sub-cohorts when comparing sub-cohorts with similar time from immunity-conferring event (Table 2), but the protection of the Vaccinated cohort can be restored by administration of the booster (Table 2).

In agreement with other studies,^6,7,21^ we found that after several months persons with hybrid immunity are better protected against further infection than uninfected persons who have received two vaccine doses (the Vaccinated cohort). Furthermore, we found that a single dose of vaccine given to a previously infected individual or to an uninfected doubly vaccinated individual (i.e., a booster dose) restores the protection to the level in the early months following recovery or vaccination. The timing of vaccination following infection affects the protection^6^. Figure S2 presents the distribution of time between infection and vaccination in our database. However, we did not have enough data to evaluate the level of protection as a function of time between infection and vaccination while taking the waning effect into account.

Our results are in line with those of a study conducted by an Israeli HMO,^7^ that previously infected individuals with or without one vaccination dose have better protection than uninfected doubly-vaccinated individuals 3 to 8 months after the last immunity-conferring event. Our data on Covid-19 hospitalized patients with severe disease has too few cases for a definitive analysis but does not seem to support a recent report^22^ that suggests that vaccinated individuals were more protected than previously infected individuals 3 to 6 months after the immunity-conferring event.

The first infections of individuals in the Recovered and hybrid cohorts were from mostly the pre-Alpha and the Alpha variants.^17^ If protection provided by prior infection depends on the variant, its effect is confounded with the effect of time since infection. As a single variant was dominant in Israel during each of the pandemic waves,^17^ our study cannot disentangle the two effects. Moreover, infections during the study period were mostly of the Delta variant, and there is not enough information at this time to suggest implications from our results regarding protection from new variants such as the Omikron.

Because our results pertain to the rate of confirmed infection, they are sensitive to detection bias due to different tendencies to perform PCR testing in the study cohorts. During the study period, previously-infected individuals and doubly-vaccinated individuals had the same official PCR testing policy, which required PCR testing upon contact with an infected individual. While differences in testing rates between cohorts and between sub-cohorts within cohorts are observed (Figure S3), their overall magnitude is relatively small. As the rate of PCR testing is typically lower in the Recovered cohort, the level of protection in this cohort when compared to the vaccinated cohort may be slightly overestimated. Our data regarding severe disease do not suffer from this bias.

Another source of potential bias is due to cohort misclassification. To be classified as a recovered person in our study, a PCR test must have been performed and found positive. However, many infected individuals have not been diagnosed^23^ and some of these have received vaccination. Thus, some of those classified into the Vaccinated cohort or the Booster cohort should have been included in the hybrid-immunity cohorts. This may have led to underestimation of the rate among vaccinated uninfected individuals. Yet, as the Recovered group was much smaller than the Vaccinated group (see Table 1), the size of this bias is expected not to be large.

Understanding the waning rates after different immunity-conferring events is important for policy making regarding the need and timing of additional vaccination doses. We found that protection against the Delta variant wanes over time for both vaccinated and previously infected individuals, and that an additional dose restores protection. Future studies will help determine the optimal timing of that dose.

## Data Availability

Aggregated data and code for reproducing the results are available from the corresponding author upon request

## Ethics statement

The study was approved by the Institutional Review Board of the Sheba Medical Center. Helsinki approval number: SMC-8228-21.

