## Supplementary Appendix for "Protection and waning of natural and hybrid COVID-19 immunity"

### Supplementary Methods 1 - Description of the Data

The analysis is based on the Israel Ministry of Health's database. Israel has experienced four pandemic waves, with the Delta (B.1.617.2) variant being the predominant variant during the fourth wave. During the third wave, Israel initiated a very rapid vaccination campaign offering the BNT162b2 vaccine to all adult residents. The campaign was opened on December 20, 2020, initially to people aged 60 years or older, and was then gradually extended[1] until, on February 4, 2021, all individuals aged 16 or older were eligible to receive two doses of the vaccine. In March 2021, previously infected individuals were eligible to receive a single BNT162b2 dose at least three months after recovery from Covid-19. After the arrival of the Delta variant to Israel, a new Covid-19 wave began in mid-June 2021. Consequently, on July 30, 2021, the administration of a third (booster) dose was approved, first for people aged 60 years or older, and later for younger age groups.[2,3]

Israel has a centralized health system, where each resident belongs to one of four health maintenance organizations (HMOs). Polymerase Chain Reaction (PCR) tests for SARS-CoV-2 infections as well as vaccination against the virus are provided free of charge, and are directly reported to the Ministry of Health (MoH). The MoH established a centralized Covid-19 national database containing regularly updated information on all PCR tests and results, vaccination dates, and follow-up data on all infected individuals, including severity of disease and mortality. In this study, re-infection is defined as a positive PCR test in an individual who had a previous positive result on a sample taken at least 90 days earlier.[4] Severe disease is defined following the US NIH definition: resting respiratory rate of more than 30 breaths per minute, an oxygen saturation of less than 94% while breathing ambient air, or a ratio of partial pressure of arterial oxygen to fraction of inspired oxygen of less than 300. The MoH database also includes basic demographic information, such as sex, age, place of residency, and population sector.

### Supplementary Methods 2 - Calculation of adjusted rates

The adjusted rates were calculated as follows. After fitting the Poisson model, the expected number of cases for sub-cohort $k$ was calculated by assuming all individuals belonging to that sub-cohort. These estimates were calculated by applying the results of the Poisson model to all persons with their coefficient of sub-cohort being replaced with that of sub-cohort $k$: $exp(\gamma_k + \beta \times covariates)$, where $exp$ is the exponential function, $\gamma_k$ is the coefficient of sub-cohort $k$, and $\beta$ is the row vector of coefficients for all other covariates (see Table S5). These estimates (expressed per 100,000 person days) are reported as the adjusted rates.

*Table S1: Demographic and clinical characteristics of the different Recovered sub-cohorts.The table presents the proportion of person-days at risk and the number of events that were used in the analysis; study period: August 1, 2021, to September 30, 2021.*

| Group | Recovered 4-6 months ago *Person-days at risk = 1,264,699* | | | Recovered, 6-8 months ago *Person-days at risk = 7,210,192* | | | Recovered 8-10 months ago *Person-days at risk = 3,243,005* | | | Recovered 10-12 months ago *Person-days at risk = 3,262,444* | | | Recovered 12+ months ago *Person-days at risk = 2,900,655* | | |
|---|---|---|---|---|---|---|---|---|---|---|---|---|---|---|---|
| | *% person days at risk* | *# Infect-ions* | *# severe Covid-19* | *% person days at risk* | *# Infect-ions* | *# severe Covid-19* | *% person days at risk* | *# Infect-ions* | *# severe Covid-19* | *% person days at risk* | *# Infect-ions* | *# severe Covid-19* | *% person days at risk* | *# Infect-ions* | *# severe Covid-19* |
| *Female* | *57.8%* | *69* | *0* | *55.6%* | *681* | *2* | *55.3%* | *455* | *2* | *47.9%* | *599* | *1* | *48.1%* | *566* | *4* |
| *Male* | *42.2%* | *52* | *2* | *44.4%* | *543* | *7* | *44.7%* | *292* | *2* | *52.1%* | *598* | *4* | *51.9%* | *536* | *1* |
| *Age 16-39* | *67.1%* | *104* | *0* | *68.2%* | *989* | *3* | *64.0%* | *551* | *3* | *66.3%* | *915* | *0* | *66.1%* | *802* | *0* |
| *Age 40-59* | *24.4%* | *13* | *0* | *24.4%* | *208* | *3* | *27.0%* | *180* | *1* | *24.7%* | *249* | *3* | *25.1%* | *265* | *3* |
| *Age 60+* | *8.6%* | *4* | *2* | *7.3%* | *27* | *3* | *9.0%* | *16* | *0* | *9.1%* | *33* | *2* | *8.8%* | *35* | *2* |
| *General Jewish* | *59.3%* | *94* | *2* | *55.4%* | *789* | *5* | *45.6%* | *415* | *1* | *48.3%* | *679* | *2* | *43.7%* | *545* | *2* |
| *Arab* | *11.0%* | *16* | *0* | *25.8%* | *335* | *2* | *26.2%* | *195* | *1* | *35.2%* | *466* | *1* | *37.3%* | *451* | *0* |
| *Ultra-Orthodox* | *29.7%* | *11* | *0* | *18.7%* | *100* | *2* | *28.2%* | *137* | *2* | *16.6%* | *52* | *2* | *19.0%* | *106* | *3* |

*Table S2: Demographic and clinical characteristics of the different Vaccinated and Booster sub-cohorts.The table presents the proportion of person-days at risk and number of events that were used in the analysis; study period: August 1, 2021, to September 30, 2021.*

| Group | Twice Vaccinated 0-2 months ago *Person-days at risk = 4,889,023* | | | Twice Vaccinated 2-4 months ago *Person-days at risk = 9,247,416* | | | Twice Vaccinated 4-6 months ago *Person-days at risk = 98,156,329* | | | Twice Vaccinated 6-8 months ago *Person-days at risk = 71,921,540* | | | Booster *Person-days at risk = 80,428,946* | | |
|---|---|---|---|---|---|---|---|---|---|---|---|---|---|---|---|
| | % person days at risk | # Infect-ions | # severe Covid -19 | % person days at risk | # Infect-ions | # severe Covid -19 | % person days at risk | # Infect-ions | # severe Covid-19 | % person days at risk | # Infect-ions | # severe Covid -19 | % person days at risk | # Infect-ions | # severe Covid-19 |
| Female | 51.1% | 689 | 9 | 52.4% | 2,405 | 24 | 50.9% | 39,876 | 158 | 51.4% | 34,505 | 388 | 51.0% | 2,835 | 70 |
| Male | 48.9% | 460 | 12 | 47.6% | 1,497 | 19 | 49.1% | 30,209 | 212 | 48.6% | 30,837 | 550 | 49.0% | 3,010 | 108 |
| Age 16-39 | 79.5% | 913 | 0 | 68.1% | 2,901 | 1 | 63.8% | 48,182 | 16 | 44.0% | 32,475 | 27 | 16.4% | 1,156 | 1 |
| Age 40-59 | 13.7% | 138 | 5 | 22.0% | 735 | 10 | 28.5% | 18,287 | 80 | 36.0% | 23,665 | 169 | 30.1% | 2,042 | 13 |
| Age 60+ | 6.8% | 98 | 16 | 9.9% | 266 | 32 | 7.8% | 3,616 | 274 | 20.0% | 9,202 | 742 | 53.4% | 2,647 | 164 |
| General Jewish | 75.6% | 887 | 18 | 70.5% | 3,149 | 32 | 71.8% | 56,494 | 267 | 77.7% | 50,644 | 754 | 89.2% | 4,957 | 155 |
| Arab | 6.7% | 104 | 2 | 6.6% | 363 | 1 | 5.8% | 6,104 | 24 | 5.1% | 5,757 | 47 | 4.1% | 417 | 12 |
| Ultra-Orthodox | 17.8% | 158 | 1 | 22.9% | 390 | 10 | 22.4% | 7,487 | 79 | 17.2% | 8,941 | 137 | 6.7% | 471 | 11 |

*Table S3: Demographic and clinical characteristics of the different Recovered then Vaccinated sub-cohorts.The table presents the proportion of person-days at risk and number of events that were used in the analysis; study period: August 1, 2021, to September 30, 2021.*

| Group | Recovered then Vacc 0-2 months ago *Person-days at risk = 2,321,324* | | | Recovered then Vacc 2-4 months ago *Person-days at risk = 2,064,746* | | | Recovered then Vacc 4-6 months ago *Person-days at risk = 3,979,206* | | | Recovered then Vacc 6-8 months ago *Person-days at risk = 1,304,879* | | |
|---|---|---|---|---|---|---|---|---|---|---|---|---|
| | % person days at risk | # Infect-ions | # severe Covid -19 | % person days at risk | # Infect-ions | # severe Covid -19 | % person days at risk | # nfect-ions | # severe Covid -19 | % person days at risk | # Infect-ions | # severe Covid -19 |
| Female | 53.3% | 61 | 0 | 54.5% | 52 | 1 | 50.8% | 245 | 7 | 47.0% | 83 | 0 |
| Male | 46.7% | 44 | 1 | 45.5% | 42 | 1 | 49.2% | 219 | 0 | 53.0% | 76 | 3 |
| Age 16-39 | 65.7% | 79 | 0 | 59.8% | 69 | 0 | 52.2% | 295 | 0 | 53.0% | 100 | 1 |
| Age 40-59 | 25.1% | 19 | 1 | 28.8% | 15 | 0 | 31.1% | 126 | 3 | 30.4% | 49 | 1 |
| Age 60+ | 9.1% | 7 | 0 | 11.5% | 10 | 2 | 16.7% | 43 | 4 | 16.6% | 10 | 1 |
| General Jewish | 54.6% | 62 | 0 | 60.4% | 68 | 1 | 50.7% | 299 | 5 | 55.5% | 110 | 2 |
| Arab | 25.8% | 23 | 0 | 20.4% | 18 | 1 | 21.0% | 103 | 1 | 21.3% | 26 | 0 |
| Ultra-Orthodox | 19.7% | 20 | 1 | 19.2% | 8 | 0 | 28.2% | 62 | 1 | 23.2% | 23 | 1 |

*Table S4: Demographic and clinical characteristics of the different Vaccinated then Recovered sub-cohorts.The table presents the proportion of person-days at risk and number of events that were used in the analysis; study period: August 1, 2021, to September 30, 2021.*

| Group | Vacc then Recovered 4-6 months ago Person-days at risk = \|542,250 | | | Vacc then Recovered 6-8 months ago Person-days at risk = 1,426,582 | | |
|---|---|---|---|---|---|---|
| | *% person days at risk* | *# Infect- ions* | *# severe Covid-19* | *% person days at risk* | *# Infect- ions* | *# severe Covid-19* |
| *Female* | *54.5%* | *42* | *0* | *50.8%* | *135* | *2* |
| *Male* | *45.5%* | *23* | *0* | *49.2%* | *143* | *3* |
| *Age 16-39* | *51.8%* | *44* | *0* | *40.7%* | *153* | *0* |
| *Age 40-59* | *32.3%* | *19* | *0* | *34.8%* | *87* | *0* |
| *Age 60+* | *15.9%* | *2* | *0* | *24.5%* | *38* | *5* |
| *General Jewish* | *64.4%* | *47* | *0* | *66.5%* | *208* | *5* |
| *Arab* | *7.6%* | *9* | *0* | *15.7%* | *41* | *0* |
| *Ultra-Orthodox* | *28.0%* | *9* | *0* | *17.8%* | *29* | *0* |

*Table S5. Coefficients of Poisson regression for confirmed infection.*

| term | estimate | std.error |
|---|---|---|
| (Intercept) | -9.791 | 0.035 |
| Age group: 40-59 | -0.101 | 0.006 |
| Age group: 60+ | -0.396 | 0.009 |
| Sex: male | -0.152 | 0.005 |
| Sector: ultra-Orthodox Jewish | 0.356 | 0.009 |
| Sector: Arab | -0.324 | 0.008 |
| Exposure risk: 2nd decile (2.17,3.21] | 0.730 | 0.021 |
| Exposure risk: 3rd decile (3.21,4.22] | 0.946 | 0.020 |
| Exposure risk: 4th decile (4.22,5.19] | 1.063 | 0.020 |
| Exposure risk: 5th decile (5.19,6.2] | 1.211 | 0.020 |
| Exposure risk: 6th decile (6.2,7.35] | 1.380 | 0.019 |
| Exposure risk: 7th decile (7.35,8.64] | 1.467 | 0.019 |
| Exposure risk: 8th decile (8.64,10.3] | 1.588 | 0.019 |
| Exposure risk: 9th decile (10.3,12.6] | 1.798 | 0.019 |
| Exposure risk: 10th decile (12.6,7.15e+03] | 2.077 | 0.019 |
| Week 2 | 0.172 | 0.012 |
| Week 3 | 0.204 | 0.012 |
| Week 4 | 0.256 | 0.012 |
| Week 5 | 0.266 | 0.013 |
| Week 6 | 0.168 | 0.013 |

| | | |
|---|---|---|
| Week 7 | 0.046 | 0.014 |
| Week 8 | 0.064 | 0.014 |
| Week 9 | 0.067 | 0.017 |
| Cohort: Recovered 12+ | 0.359 | 0.042 |
| Cohort: Recovered 10-12 | 0.299 | 0.041 |
| Cohort: Recovered 8-10 | -0.026 | 0.047 |
| Cohort: Recovered 6-8 | -0.414 | 0.041 |
| Cohort: Recovered 4-6 | -0.702 | 0.096 |
| Cohort: Booster 0-2 | -0.941 | 0.033 |
| Cohort: Vaccinated 6-8 | 1.437 | 0.030 |
| Cohort: Vaccinated 4-6 | 1.187 | 0.030 |
| Cohort: Vaccinated 2-4 | 0.756 | 0.034 |
| Cohort: Recovered then Vaccinated 6-8 | -0.599 | 0.085 |
| Cohort: Recovered then Vaccinated 4-6 | -0.715 | 0.055 |
| Cohort: Recovered then Vaccinated 2-4 | -1.603 | 0.107 |
| Cohort: Recovered then Vaccinated 0-2 | -1.733 | 0.102 |
| Cohort: Vaccinated then Recovered 6-8 | -0.208 | 0.067 |
| Cohort: Vaccinated then Recovered 4-6 | -0.504 | 0.127 |

*Table S6: Summary of the results regarding confirmed infections of the Poisson regression analysis for all sub-cohorts by age groups. For each group, the table shows the estimated covariate-adjusted confirmed infection rate per 100,000 person-days at risk. 95% confidence intervals without adjustment for multiplicity are given in square brackets.*

| *Cohort* | *16-39* | *40-59* | *60+* |
|---|---|---|---|
| *Recovered 4-6* | *14.2 [11.5, 17.4]* | *5.0 [3.0, 8.7]* | *4.2 [1.6, 10.5]* |
| *Recovered 6-8* | *17.4 [16.4, 18.5]* | *10.7 [9.4, 12.3]* | *4.8 [3.3, 6.9]* |
| *Recovered 8-10* | *24.6 [22.6, 26.9]* | *20.2 [17.6, 23.3]* | *5.6 [3.4, 9.3]* |
| *Recovered 10-12* | *34.5 [32.4, 36.9]* | *26.1 [23.0, 29.5]* | *9.7 [6.8, 13.8]* |
| *Recovered 12+* | *34.9 [32.5, 37.3]* | *31.5 [28.0, 35.5]* | *12.4 [8.9, 17.2]* |
| *Booster 0-2* | *10.7 [10.1, 11.4]* | *8.9 [8.5, 9.3]* | *5.6 [5.4, 5.8]* |
| *Vaccinated 0-2* | *23.0 [21.5, 24.5]* | *19.9 [16.9, 23.5]* | *26.5 [21.6, 32.1]* |
| *Vaccinated 2-4* | *52.9 [51.2, 55]* | *40.6 [37.9, 43.8]* | *29.3 [26.2, 33.0]* |
| *Vaccinated 4-6* | *77.5 [76.8, 78.3]* | *71.2 [70.2, 72.3]* | *50.1 [48.5, 51.8]* |
| *Vaccinated 6-8* | *98.1 [97.0, 99.2]* | *89.7 [88.6, 90.8]* | *72.2 [70.8, 73.7]* |
| *Recovered then Vaccinated 0-2* | *4.5 [3.6, 5.5]* | *2.9 [1.8, 4.5]* | *3.0 [1.4, 6.1]* |
| *Recovered then Vaccinated 2-4* | *5.4 [4.3, 6.8]* | *2.5 [1.5, 4.2]* | *4.3 [2.3, 8.0]* |
| *Recovered then Vaccinated 4-6* | *12.6 [11.2, 14.1]* | *9.6 [8.0, 11.4]* | *6.0 [4.5, 7.9]* |
| *Recovered then Vaccinated 6-8* | *13.9 [11.5, 16.7]* | *12.3 [9.3, 16.3]* | *4.7 [2.5, 8.8]* |
| *Vaccinated then Recovered 4-6* | *17.3 [12.9, 23.1]* | *12.0 [7.6, 18.3]* | *2.4 [0.6, 9.5]* |
| *Vaccinated then Recovered 6-8* | *22.9 [19.5, 26.9]* | *15.3 [12.4, 18.6]* | *9.7 [7.0, 13.2]* |

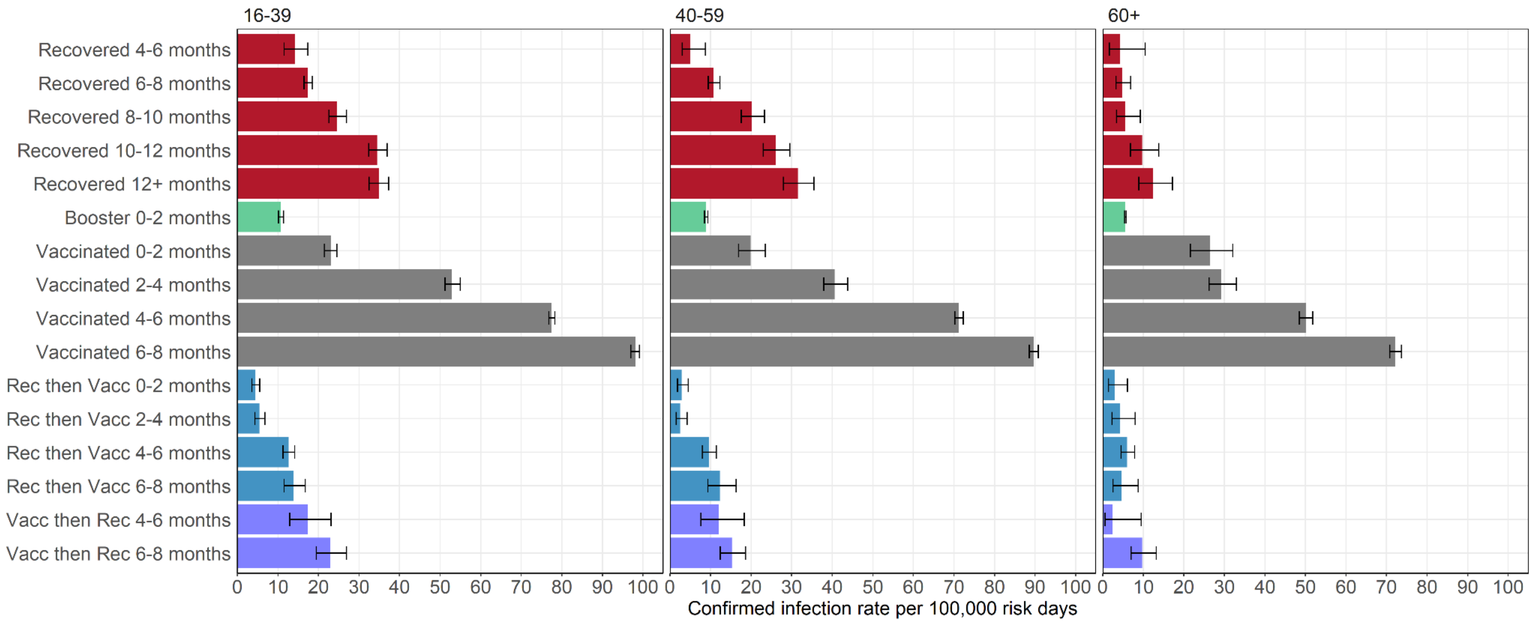

*Figure S1: Estimated covariate-adjusted rates of confirmed infections per 100,000 at-risk days obtained from the Poisson regression analysis for the study period August 1, 2021, to September 30, 2021, stratified by age and sub-cohorts. Confidence intervals are not adjusted for multiplicity.*

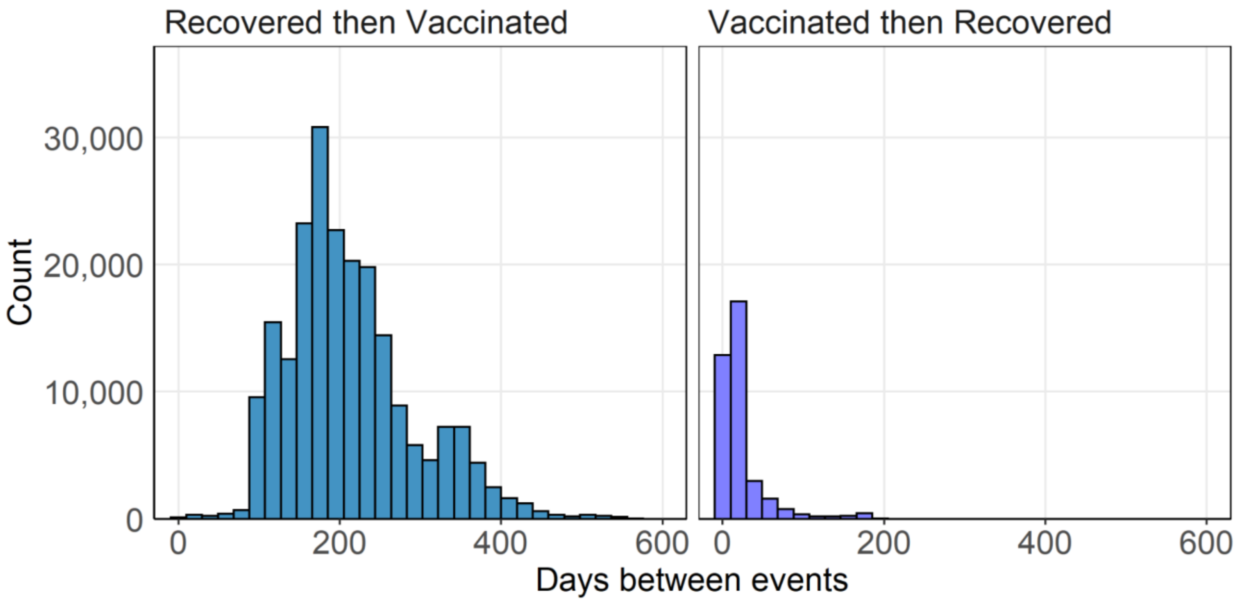

*Figure S2: The distribution of time between infection and vaccination (left) in the Recovered then Vaccinated cohort, and between vaccination and infection (right) in the Vaccinated then Recovered cohort. The latter is shorter as individuals become doubly vaccinated starting January 2021.*

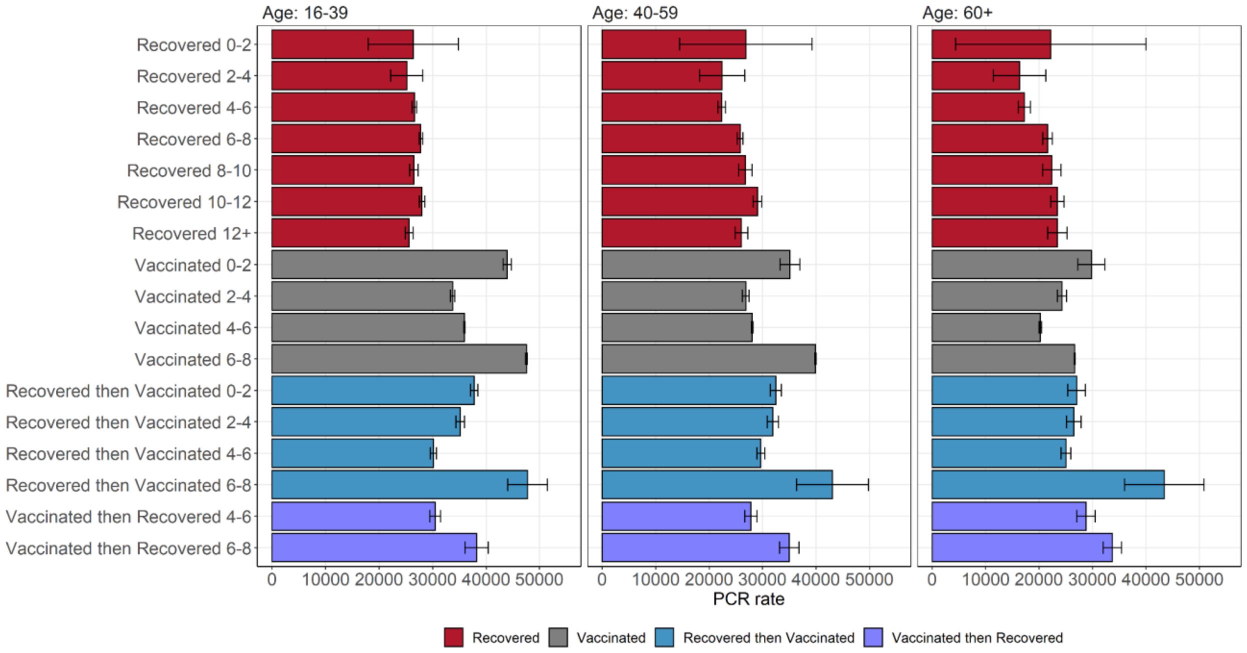

*Figure S3: PCR testing rates by sub-cohort and age group. Bars indicate the number of individuals, per 100,000, who performed at least one test during the study period. Individuals were associated with their sub-cohort at the beginning of the study.*
